# Towards molecular stratification of pediatric T-cell lymphoblastic lymphomas based on Minimal Disseminated Disease and *NOTCH1/FBXW7* mutational status: the French EURO-LB02 experience

**DOI:** 10.1101/2020.09.08.20189829

**Authors:** Amélie Trinquand, Adriana Plesa, Chrystelle Abdo, Nathalie Aladjidi, Charlotte Rigaud, Aurore Touzart, Ludovic Lhermitte, Arnaud Petit, Katell Michaux, Charlotte Jung, Catherine Chassagne-Clement, Vahid Asnafi, Yves Bertrand, Nathalie Garnier, Elizabeth Macintyre

## Abstract

While outcome for pediatric T lymphoblastic lymphoma (T-LBL) has improved with Acute Leukemia-type therapy, survival after relapse remains rare. Few prognostic markers have been identified and the value of Minimal Residual Disease (MRD) is less clear than in T-ALL. Mutations of *NOTCH1* and/or *FBXW7* (*N/F*) identify good prognosis T-LBL and both MRD and high-level Minimal Disseminated Disease (MDD) are reported to be of poor prognosis. We evaluated MDD status by 8-color flow cytometry (MFC) and/or digital droplet PCR (ddPCR) in 86 French pediatric T-LBL, of which *N/F* status was known for 65 (61 treated on the Euro-LB02 protocol). Both techniques gave identical results for MDD/MRD values above 0.1%, allowing compilation. While an MDD threshold of 1% had no prognostic significance, the 54% (44/82) of protocol-treated patients with MDD ≥0.1% had a relatively favorable outcome (overall survival/OS; p = 0.026). MDD 0.1% status had no prognostic significance in the 68% of patients with *N/F* mutations, whereas low/negative MDD status (9/61) identified *N/F* germline patients at a high risk of relapse (5-year OS of 44.4% vs 90% for MDD ≥ 0.1%,p = 0.014; and a 5-year DFS of 50% vs 90.9% respectively, p = 0.041). Combining oncogenetic and MDD status allows identification of 85% of patients with an excellent outcome (5-year OS 91.9% and DFS 95%) and 15% of *N/F* germline/MDD< 0.1% patients who clearly require early alternative treatment (5-year OS 44.4%; p< 0.0001 and DFS 50%; p = 0.0001).

## INTRODUCTION

T-cell lymphoblastic lymphoma (T-LBL) and T-cell acute lymphoblastic leukemias (T-ALL) occur predominantly in children and adolescents. They are both characterized by the proliferation of malignant immature T-cell precursors but differ by the extent of bone marrow (BM) involvement, which is (arbitrarily) less than 25%, assessed by morphology, in T-LBL. Despite the improvement of current therapy, achieving an Event-Free Survival (EFS) at 5 years from 75% to 85%^1–3^, the survival rate of refractory or relapsed lymphoblastic lymphomas remains very poor, at 10-30%^4,5^. Relatively favourable results are obtained in the rare T-LBL cases who achieve second remission and can undergo hematopoietic stem cell transplantation (HSCT)^6^. The early identification of poor risk T-LBL is thus mandatory.

Contrary to T-ALL^7,8^, prognostic factors in T-LBL are not clearly established, partly due to its low incidence and difficulty in obtaining diagnostic material. Retrospective analyses identified the absence of *NOTCH1* and/or *FBXW7* (*N/F*) mutations and biallelic T-cell receptor-gamma (TRG) deletions to be associated to unfavourable outcome^9^ and loss of heterozygosity at chromosome 6q (LOH6q) to an increased relapse risk^10^.

In T-ALL, one of the strongest prognostic factor is minimal residual disease (MRD)^8^. Unlike B lineage ALL, MRD analysis in peripheral blood (PB) and BM give comparable results^11^. In T-LBL, it has been reported that evaluation of minimal disseminated disease (MDD) at diagnosis and MRD by flow cytometry and real-time quantitative polymerase chain reaction (qPCR) for TR rearrangements allows identification of high risk patients^12–14^. In these studies, MDD detection superior to 3-5% in PB or BM were associated with unfavourable EFS^12,13^. qPCR is a well-established tool for MRD detection, notably in leukemia, but has limitations in lymphoma because of the need for a reference standard curve, based on tumour-specific target serial dilutions from diagnostic DNA with known infiltration, usually assessed by flow cytometry (FC)^15^. Evaluating tumour infiltration is more difficult in diagnostic tissue. Droplet digital PCR (ddPCR) is based on sample compartmentalization in single oil droplets. An independent end-point PCR reaction occurs in each droplet, permitting absolute quantification of target DNA molecules using Poisson statistics. Studies indicate that ASO-specific ddPCR is particularly adapted for mature B cell lymphomas^16–18^ but ddPCR has not, to date, been evaluated in T-LBL.

In the French EURO-LB02 cohort, a minority of patients benefitted from prospective MDD and MRD evaluation by 8-color FC. The present study compared and extended these results with TR ddPCR analyses in order to assess the clinical impact of MDD at diagnosis. Since the recently opened European multicentre prospective pediatric LBL2018 protocol (NCT04043494) stratifies patients on their *N/F* mutation status, we also evaluated the correlation between these oncogenetic markers and MDD on outcome. We showed that, unexpectedly, patients with MDD positivity above 0.1% in PB/BM have a relatively favourable prognosis, essentially within the poor prognosis *N/F* germline (*N/F*^GL^) subgroup.

## PATIENTS AND METHODS

### Patients

The EURO-LB02 protocol was a randomized controlled study, derived from the Berlin-Frankfurt-Munich BFM90 protocol but without prophylactic central nervous system irradiation, involving patients aged < 22y with *de-novo* LBL in 8 European cooperative groups (NCT00275106)^19^. The study was interrupted prematurely due to excessive toxicity, but the prednisone arm was continued in France, with central registration of clinical data in Lyon. From 2008 onwards, prospective flow cytometry was performed centrally (Lyon) for the French cohort. Between December 2004 and December 2017, 299 patients with T-LBL were enrolled in 30 institutions of the Société Française de lutte contre les Cancers et leucémies de l’Enfant et de l’adolescent (SFCE). As per the declaration of Helsinki, the Lyon ethics committee approved the study and signed informed consent was obtained for all patients. Patients who received systemic corticosteroids for more than 8 days within 2 months before or at the time of MDD evaluation were excluded from this study, although lower dose oral corticotherapy cannot be excluded for all patients. The present cohort of 82 patients did not differ from the overall French cohort. Two patients died in remission of toxic complications. Eighteen patients diagnosed between 2004-2010 were included in prior description of *N/F* status^9^

### MDD/MRD assessment by Multiparameter Flow Cytometry

From 2008 to 2017, fresh PB and/or BM samples were collected at diagnosis and PB at day (d)33 (end of induction). In total, 109 MDD samples were analysed in 66 patients and 58 patients had 67 MRD samples (46 PB only, 3 BM only, 9 both). Multiparameter flow cytometry (MFC) was performed with an 8-color flow panel on a FACS CANTO II with DIVA software (BD Bioscience), as described^20,21^. Instrument set-up was regularly calibrated using Calibrite, Rainbows 8 picks, and CST beads systems. Positive MDD/MRD were defined as a cluster of more than 10 cells expressing at least two LAIP (Leukemia Aberrant Immuno-Phenotype) and SSC characteristics identified at diagnosis, amongst at least 500 000 viable cells. MFC sensitivity of 1×10^−4^ (0.01%) normal cells was possible in virtually all samples. Whenever possible, leftover cells were frozen as a pellet for molecular quantification.

### MDD/MRD assessment by Droplet Digital PCR (ddPCR)

44 patients with fresh or cryopreserved MDD samples (40 BM and 20 PB) and 15 patients with d33 MRD frozen samples (5 BM and 14 PB) were analysed retrospectively (Paris). To identify patient-specific clonotypic markers, genomic DNA was extracted from diagnostic tissue or effusions using a QIAampDNA mini kit (Qiagen Co., Hilden, Germany), TR clonality (TRD, TRB and TRG) was assessed by one-step Next-Generation Sequencing using 100ng DNA, EuroClonality-NGS amplicon primers^22^ and Vidjil software analysis^23^. At least one allele-specific (ASO) CDR3 primer, selected on the basis of maximal clonotype amplification and minimal non-specific positivity in normal PBL DNA, was designed for each patient.

Quantification of MDD and MRD was performed by ddPCR with the QX100 Droplet Digital PCR system (Bio-Rad Laboratories, Hercules, CA). 250ng (5µl) gDNA were amplified in 20µl with 10µl of 2X ddPCR Master Mix (Bio-Rad Laboratories), 1µl of 20X primers and probe (final concentration, 500nmol/l and 200nmol/l). End point PCR was performed on a T100 Thermal Cycler (Bio-Rad Laboratories) followed by droplet analysis on the QX100 reader using QuantaSoft V1.2 software, as per the manufacturer’s recommendations. Each experiment included 2 replicates with no template control (NTC), a positive control using 100ng of diagnostic gDNA, 3 replicates of 250ng of each MDD/MRD sample, 2 replicates of 250ng of a 0.01%/10^−4^ dilution of diagnostic DNA (corrected for initial infiltration) to evaluate sensitivity and 3 replicates of 250ng pooled PBMC gDNA from 6 healthy donors, to assess non-specific ASO amplification. The final tumour load was calculated as a mean of all technically acceptable replicates after application of a Poisson correction and exclusion of nonspecific channel 2 false positivity. DNA quantity and amplifiability was assessed in a separate reaction by quantification of the albumin housekeeping gene from 100ng of gDNA sample in a single well, total volume 20μl.

ddPCR results were analysed using updated criteria defined for Mantle Cell Lymphoma^17^. Briefly, only replicates with ≥9000 droplets were accepted and a single threshold for all samples above the highest background value and below the positive control was set. Positive samples were those with a merge of events ≥3 regardless of the number of positive triplicates. Below quantitative level (BQL) samples had a merge of events of 2. Undetectable samples had, at most, replicates with only one event. If there was a merge of ≥2 events in 3 PBMC with ≥9000 droplets, we subtracted the number of non-specific events in the PBMC from the total number of events in the sample, as described^17^. Target results were corrected for Albumin quantitation, since the majority of MDD samples were frozen samples leftover after MFC analysis or fresh/cryopreserved or FFPE tissue samples (n = 2).

### *NOTCH1/FBXW7* mutations

The mutational screening of tumour samples with at least 20% infiltration was performed by capture NGS, as previously described^24^.

### Statistical Analysis

The association between presenting features and presence of T-LBL cells by flow cytometry was analysed with the Chi2, the Student test or Pearson correlation test. The log-rank test was used in the Kaplan-Meier estimations of Disease-Free Survival (DFS) and overall survival (OS) with Prism 6 software (GraphPad Software Inc.).

## RESULTS

### Minimal Disseminated Disease assessment by Multiparametric Flow Cytometry

From 2008 to 2017, 66 pediatric T-LBL had MDD quantification by 8-colour MFC from 109 samples (21 PB only, 2 BM only, 43 both) (Figure S1). Among them, 62 patients were included/treated according to EURO-LB02. As described^20,21^, optimal combinations to measure MRD in T-ALL were those containing TdT, CD99, CD1a and CD34, in addition to a CD3/cCD3/CD5 backbone. To exclude residual erythrocytes, dead cells, debris, platelet aggregates and doublets, a live gate was adjusted on forward scatter (FSC)/SSC and CD45/SSC. It was possible to compare pathological MDD phenotypes with those of the tumour in only 9 patients for whom a pleural or pericardial effusion was transmitted for central MFC. Despite this, a pathological population of at least 0.01% was identified in 81/109 PB/BM samples (53/66 patients), with a mean infiltration of 1.8% (range 0.01-15%), when analysing at least 500 000 viable cells. (Figure S2A, B). Infiltration was similar in peripheral blood and bone marrow in the 43 patients with dual analysis (data not shown).

### MDD quantitation by ddPCR

In order to evaluate ddPCR MDD/MRD absolute quantitation, TR immunogenetic status was evaluated in diagnostic DNA from 66 infiltrated tumour samples with known MDD/MRD status (by MFC and/or ddPCR) by genescan multiplex clonality analysis^25^ and/or EuroClonality amplicon NGS for TRD, TRG and TRB (VDJ and DJ) rearrangements^22^. Clonal rearrangement of at least one TR was identified in all but one case, for which MDD analysis was performed by MFC, with TRG clonality identified in 92.4% (61/66), TRB VDJ+/−DJ in 84.8% (56/66), TRB DJ only in 7.6% (5/66), and TRD in 48.4% (31/64). At least one ASO CDR3 primer was developed for each of the 48 T-LBL with MDD/MRD samples and first tested by ddPCR on diagnostic tissue samples. Infiltration in the 27 pleural/pericardiac effusions varied from 4-100%, median 46% and from 40-90%, median 56%, in tissue samples. As such, diagnostic liquid and tissue samples cannot be presumed to always be massively infiltrated at the DNA level, regardless of the histological interpretation.

For MDD ddPCR quantification, we first evaluated cell pellets frozen after prospective MFC analysis. The sensitivity of the selected ASO was at least 0.01% in all cases. Since the quantity of DNA available from the cell pellets was often limited, we compared quantification from triplicates of 250ng (112500 cell equivalents) to 500ng triplicates (225000 cell equivalents) on 10 positive samples, but there was no obvious increased sensitivity with the latter (data not shown) so 250ng triplicates were retained. Target quantification was corrected for DNA quantity and quality, based on albumin quantification in a separate reaction^17^, since the majority of MDD samples were frozen pellets leftover from MFC analysis.

### ddPCR comparison with MFC

A total of 65 samples (PB, BM or effusions at diagnosis or d33) from 29 patients were evaluated by both methods Fig. 1A). MFC and ddPCR showed excellent overall concordance (Pearson correlation test: r = 0.829 and p< 0.0001). There was absolute concordance above the threshold of 0.1%, but ddPCR gave quantifiable positivity in 10/30 cases (plus one BQL result) with no detectable infiltration by MCF, in keeping with reported sensitivity limits of 0.01% for MFC, compared to 0.001% by ddPCR, at least in MCL^17^. For 6 samples (3 patients) with ddPCR quantification between 0.1-0.01%, MFC raw data review confirmed the absence of detectable T-lymphoblasts, albeit in patients whose tumour immunophenotype had not been assessed. In 5 samples (3 MDD), ddPCR was positive/BQL below the sensitivity of MFC (0.01%). Taken together, these results demonstrate that ddPCR is a sensitive method for MDD/MRD assessment in T-LBL, that MFC and ddPCR can be compiled for values above 0.1% and that ddPCR is more sensitive for low level positivity than MCF, as practised here (0.5-1 million cells analysed).

**Figure 1:**
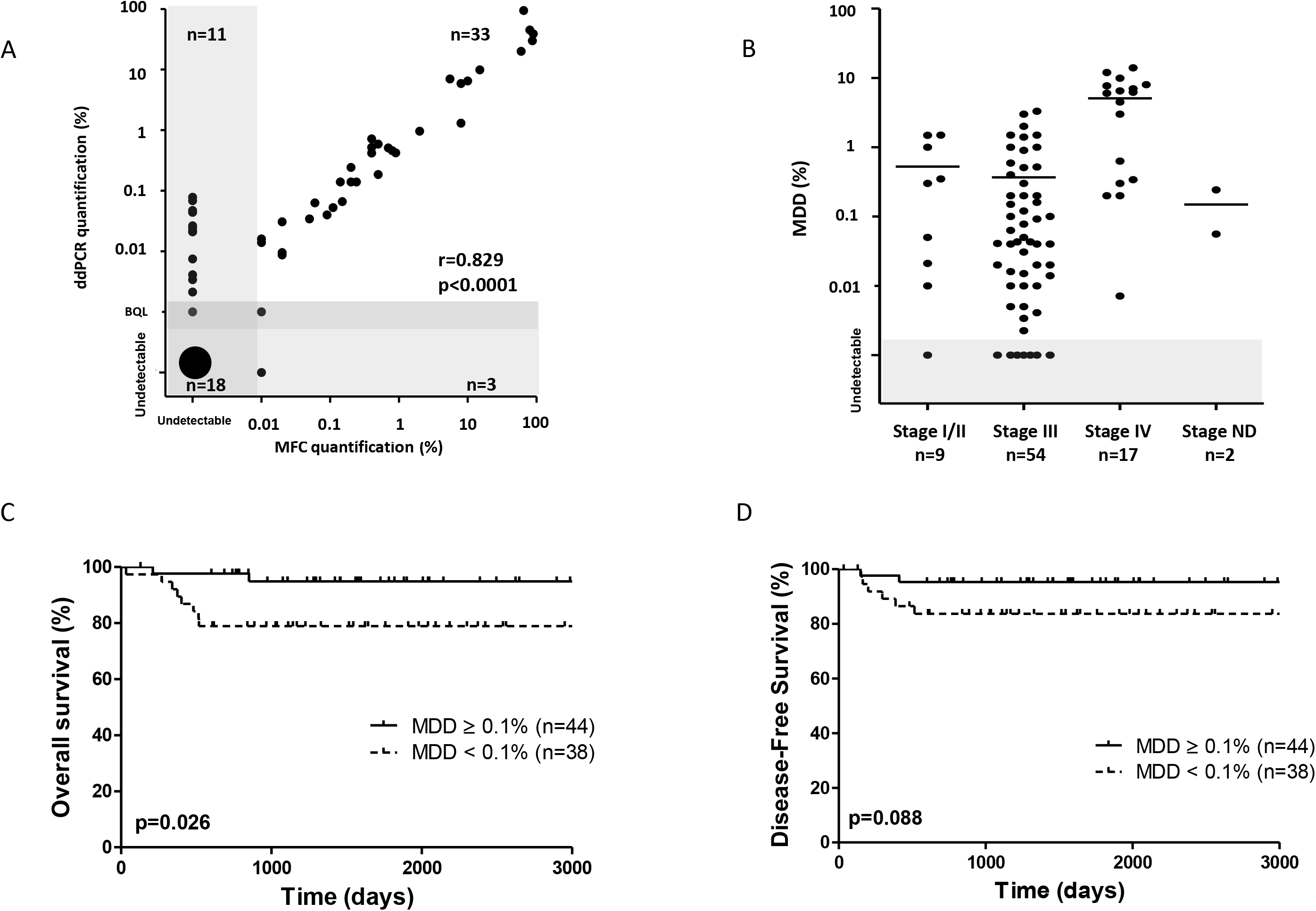
Minimal Disseminated Disease in pediatric T-LBL. (A) MFC and ddPCR gave comparable quantitative results. 65 pleural effusions, PB and BM samples at diagnosis or follow-up were analysed by both methods (Pearson r = 0.829 p< 0.0001, R^2^: 0.6878). (B) MDD quantification according to clinical stage. (C) Overall Survival with a MDD 0.1% threshold. (D) Disease-Free Survival with a MDD 0.1% threshold.

### MDD infiltration at diagnosis

We extended our ddPCR study to all frozen PB/BM MDD/MRD samples for which a tumour sample with a clonal TR was also available (Figure S2B). In total, 48 patients were evaluated by ddPCR, including 44 with MDD quantification. ASO targets were TRB for 31 patients, TRD for 10 and TRG for 7. Only 2 patients had enough DNA to be quantified on 2 targets, when the most positive result was retained. All samples met the defined ddPCR criteria^17^, (sufficient replicates and droplets). The vast majority (41/44, 93%) of samples were quantifiably positive, with 7 samples in the 0.001-0.01% range and only 1 positive BQL (Figure S2B).

When combined with MFC results, 86 patients had MDD evaluation by MFC, ddPCR or both (Fig. S2B), including 82 EURO-LB02 registry patients. For each patient, the highest MDD value was retained, regardless of sample type or assay used. We observed a continuum of infiltration in PB/BM, with 9% (8/82; 5 by MFC only, 3 by ddPCR +/− MFC) of patients having undetectable disease, 36% (30/82) positivity < 0.1%, 27% (22/82) positive ≥ 0.1% and < 1% and 27% (22/82) ≥1%. Overall, 54% of patients were MDD positive above 0.1%, at which level MCF and ddPCR were totally concordant.

### MDD ≥0.1% identifies patients with a good respond to standard EUROLB02 therapy

In order to evaluate the prognostic relevance of MDD status within the French LB02 cohort, we first verified that the patients with MDD analysis were representative of the overall cohort (Table 1). Rare studies, using an relatively high cut-off, have suggested that patients with MDD ≥ 1–5% by MFC have a poor prognosis^12,13^. Using a 1% threshold for the present series, MDD had no prognostic value (5-year Overall Survival (OS) of 90% for patients with MDD ≥ 1% versus 87% for patients with MDD < 1%, p = 0.64; and 5-year Disease-Free Survival (DFS) of respectively 91% versus 90%, p = 0.93; Fig. S2C and D). In contrast and unexpectedly, when using a MDD cut-off of 0.1%, above which there was an absolute concordance between MFC and ddPCR, patients with MDD ≥ 0.1% had a favourable 5-year OS of 95% compared to 79% for patients with MDD < 0.1%, p = 0.026, with a similar trend for 5-year DFS (95% versus 84%, p = 0.088) (Figure 1C and D).

In order to investigate whether there were clinical differences in MDD (cut-off 0.1%) high and low/neg. patients, we compared clinical and biological characteristics of the two groups (Table 1). Age, sex, CNS involvement and number of relapse were comparable. As expected, stage IV patients and/or those with BM involvement had higher MDD values (Fig. 1B).

**Table 1:** Clinical and biological characteristics of the French MDD cohort treated on EURO-LB02.

|  | French Euro-LB02 cohort<br>(n=299) | MDD cohort<br>(n=82) | MDD cut-off 0.1% |  |  |
| --- | --- | --- | --- | --- | --- |
|  |  |  | < 0.1 %<br>(n=38) | ≥ 0.1 %<br>(n=44) | p |
| **Clinical features** |  |  |  |  |  |
| Age, mean in years (range) | 9.3 (0.5-18) | 10.1 (0.5-17.7) | 10.8 (2.9-16.9) | 9.4 (0.5-17.7) | ns |
| Gender (M, %) | 226 (76%) | 63 (77%) | 28 (74%) | 35 (80%) | ns |
| Stage |  |  |  |  |  |
| I/II | 19 (7%) | 9 (11%) | 4 (11%) | 5 (12%) | ns |
| III | 213 (73%) | 54 (68%) | 32 (86%) | 22 (51%) | p=0.0008 |
| IV | 57 (20%) | 17 (21%) | 1 (3%) | 16 (37%) | p=0.0002 |
| CNS involvement (Yes, %) | 8/149 (5%) | 5/79 (6%) | 1/36 (3%) | 4/43 (9%) | ns |
| BM involvement (Yes, %) | na | 13/79 (16%) | 0 | 13/42 (31%) | p=0.0001 |
| Relapse (Yes, %) | 36/270 (13%) | 8 (10%) | 6 (10%) | 2 (5%) | ns (p=0.14) |
| **Biological features** |  |  |  |  |  |
| LDH, xN (average, range) | na | 2.4 (0.6-15.2) | 3.0 (0.6-6.2) | 1.9 (0.6-15.2) | ns |
| *NOTCH1*/*FBXW7* mutation (Yes, %) | / | 41/61 (66%) | 20/29 (69%) | 21/32 (66%) | ns |
| MRD pos/BQL | / | 7/55 (13%) | 2/25 (8%) | 5/30 (17%) | ns |

Ns: not statistically significant (p>0.05); CNS: Central Nervous System; BM: bone marrow; MRD: minimal residual disease at day 33; BQL; Below quantitative level by ddPCR; na: not available.

### The prognostic impact of MDD status is restricted to *NOTCH1/FBXW7* germline T-LBL

Since pediatric T-LBL patients with *NOTCH1*/*FBXW7* germline status (*N/F*^GL^) are randomised for intensification in the LBL2018 protocol, we determined MDD impact as a function of *N/F* status. In the present cohort, 21/86 (24%) patients were not evaluated due to absence of sufficiently infiltrated diagnostic material, 21/65 (32%) were *N/F*^GL^ and 44/65 *N/F*^mut^ (68%). The level of MDD was comparable in the 3 groups (Figure 2A). As expected, *N/F*^GL^ patients had worse outcomes (5-year OS of 66% for *N/F*^GL^ vs 93% for *N/F*^mut^, p = 0.0059, data not shown; and a 5-year DFS of 70% vs 95% respectively, p = 0.0048; Figure 2B).

Using the MDD 0.1% threshold, MDD status had no prognostic impact in *N/F*^mut^ cases (5-year OS, p = 0.57; 5-year DFS, p = 0.99). In contrast, among *N/F*^GL^ cases, 9/20 (45%) were MDD < 0.1% and had a worse prognosis (5-year OS of 44.4% vs 90% for MDD ≥1%, p = 0.014; and a 5-year DFS of 50% vs 90.9% respectively, p = 0.041, Figure 2C and D). Overall, a molecular classifier combining MDD assessment and *N/F* genotype identified striking differences in outcome between high-risk patients (*N/F*^GL^ and MDD < 0.1%; 15%) and low-risk patients (all others) with 5-year OS at 44.4%; 95% CI, 13.6%-71.9% vs. 91.9%; 95% CI, 79.7%-96.9%, p< 0.0001; and 5-year DFS of 50%; 95% CI, 15.2%-77.5% vs. 94.1% 95% CI, 82.9%-98.1% respectively, p = 0.0001 (Figure 2E and F). Taken together, these results demonstrate that combined MDD and oncogenetic evaluation at diagnosis allows the identification of approximately 15% of patients with a 5 years overall survival of only 44%, who merit alternative treatment front line, given the very poor survival of relapsed pediatric TLBL.

**Figure 2:**
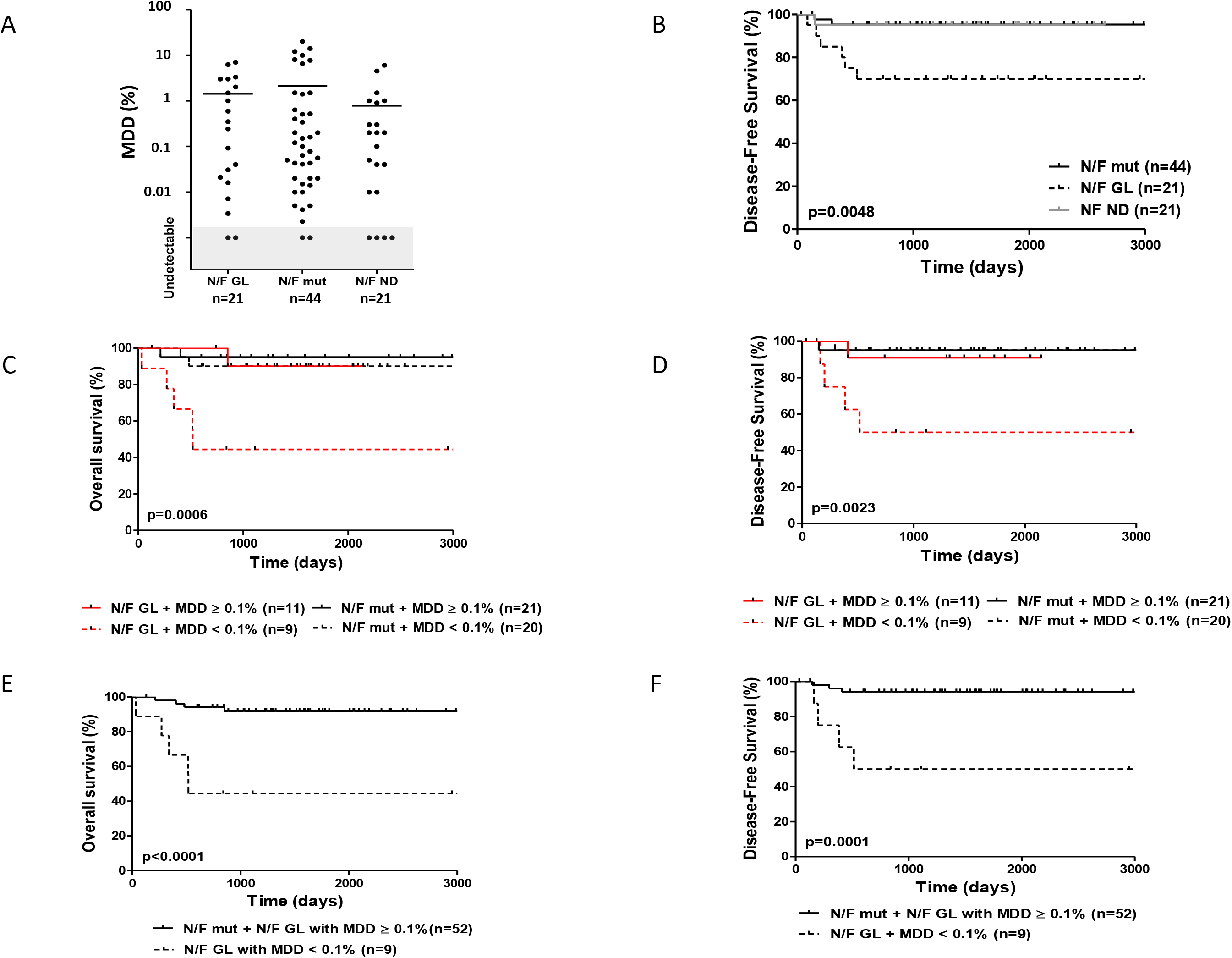
Minimal Disseminated Disease and *NOTCH1/FBXW7* mutational status in pediatric T-LBL. (A) MDD levels according to *N/F* status. (B) Disease-Free Survival according to *N/F* status. Two patients died in remission from toxicity at d33 and d482. (C) Overall Survival according to *N/F* and MDD 0.01% status. (D) Disease-Free Survival according to *N/F* and MDD 0.01% status (E) Overall Survival comparing the 15% of *N/F^GL^* and MDD< 0.01% patients with all others. (F) Disease Free Survival comparing the 15% of *N/F^GL^* and MDD< 0.01% patients with all others.

### Prognostic impact of Minimal Residual Disease

MRD at end of induction is a fundamentally important prognostic factor in T-ALL. We therefore evaluated MRD at d33 in 59 patients with available samples (41 by MFC, 17 by both CMF and ddPCR, 1by ddPCR only; Fig. S1). It is noteworthy that one *N/F^mut^* case remained persistently MRD positive by ddPCR 4 months after diagnosis, so was switched to a high risk T-ALL regimen with Nelarabine, and is in CR 3 years later. Follow-up was censured at the time of treatment intensification but this case was not considered to have relapsed. Samples from 9 patients (15%) were positive or BQL (2 by MFC+/ddPCR+, 4 ddPCR+/MFC- and 3 MFC+/ddPCR^nd^), and 50 (85%) were undetectable, with a sensitivity of 0.01%. Prognostic evaluation showed a trend for poorer prognosis in MRD positive patients, with 5-year OS of 75% for MRD positive/BQL vs 92% for MRD undetectable, p = 0.11; and a 5-year DFS of 76.2% vs 94% respectively, p = 0.061 (Figure S3). MRD alone did not predict the majority of the 9 relapses, whose clinico-biological characteristics are detailed in Table S1. The small sample size and the mixture of techniques used precludes reliable analysis of MRD significance within MDD or *N/F* defined subsets, but only 2/8 relapsing patients tested had MDD ≥0.1% and only 2/8 tested at diagnosis had *N/F^mut^* T-LBL. The data presented here suggest that this should be addressed specifically and prospectively in a collaborative study.

## DISCUSSION

T-LBL prognosis has greatly improved with ALL treatment strategies, achieving more than 85% OS. In contrast, refractory patients and those who relapse, predominantly within the first 2 years, have a dismal outcome, so their early identification is essential for timely therapeutic adaptation. We show that poor prognosis pediatric T-LBL predominates in patients with MDD levels in PB/BM below 0.1% and absence of *NOTCH1/FBWX7* mutations at diagnosis. These results contradict prior data suggesting that MDD positivity is associated with poor prognosis. We performed MDD and MRD quantification by MFC and ddPCR, following ddPCR approaches developed for Mantle Cell Lymphoma, which also frequently disseminates to PB and BM^17^. Results were perfectly concordant for positivity levels above 0.1%, allowing compilation. ddPCR was, however, more likely to detect lower level positivity, since 10/32 samples were positive/BQL at 0.01-0.1% by ddPCR, but negative by MCF, compared to only 3 which were MCF positive (all at 0.01%) but ddPCR negative. In keeping with this, a disproportionate number of positive MRD samples were detected by ddPCR. As such, either technique is acceptable for detection of high level positivity, but evaluation of levels below 0.01% should either be performed by molecular clonotype quantification or by MFC of higher cell numbers than the 0.5-1 million events analysed here. This is increasingly practised in ALL MRD evaluation, but it is noteworthy that MFC cannot be used in isolation to determine MRD negativity in the European ALL-Together trial (EUDRACT 2018-001795-38). MDD evaluation by MFC may also be challenging without characterization of the tumour phenotype and should only be assessed by reference platforms with sufficient T-LBL recruitment. ddPCR requires availability of diagnostic tissue/cells, but this is also required for oncogenetic assessment, pleading for integrated cellular and molecular oncogenetic and immunogenetic platforms.

Comparison with the largest published cohort of T-LBL (treated on COG A5971) MDD data, assessed by 9-color MFC^12^, identified the same proportion of patients with MDD ≥1% (26% vs. 27%) but fewer undetectable MDD samples (9% vs. 28%) and more patients with MDD positivity below 0.1% (36% vs. 28%), in keeping with the higher sensitivity of combined ddPCR and 8-color MFC, as performed here.

Despite the comparable incidence of high level MDD values, we did not confirm the inferior outcome of high level MDD in Euro-LB02 French cohort. On the contrary, when using a lower MDD cut-off at 0.1% (10^−3^), patients with circulating T lymphoblasts responded relatively well to ALL-type therapy. This is in keeping with the, at least, comparable outcome of Stage IV (5y EFS 88%) compared to Stage III (EFS 78%) patients in the Euro-LB02 trial^19^. Reasons for this difference are not evident, but may result from differences in treatment compared to Children’s Oncology Group Study A5971^12^. MDD low/neg. values might be due to prior steroids, since only systemic corticotherapy was an exclusion criterion, but this seems unlikely to explain the poor prognosis, since corticosensitivity is usually associated with a relatively good outcome.

Following demonstration of a relatively favourable outcome of *N/F^mut^* in pediatric T-LBL^9,10^, this parameter is now used to randomise patients for treatment intensification in the LBL2018 prospective trial. The incidence of *N/F^mut^* was slightly higher in the current cohort of patients with both oncogenetic and MDD evaluation compared to our initial assessment (66% vs. 55%)^9,^ at least partly due to the replacement of Sanger screening by next-generation-sequencing capture screening for *N/F* status^24^. The impact of MDD status was strikingly different in *N/F^mut^* and *N/F^GL^* patients. Whereas there was no prognostic impact in *N/F^mut^* patients, MDD below 0.1% clearly identified *N/F^GL^* patients at risk of relapse (p = 0.0023 for DFS and 0.0006 for OS), suggesting that significant dissemination to PB/BM may correlate with sensitivity to ALL-type therapy. Conversely, purely tissue-based disease with low/undetectable MDD negativity may be less sensitive. What this MDD cut-off should be requires further analysis with uniform techniques.

Our in-depth analysis of patient characteristics by MDD levels below 0.1% was complicated by the differential sensitivity of MFC and ddPCR. There were 8 MDD totally negative cases, of which 5 were analysed by MCF only, 3 by ddPCR +/− MCF, Table S1. They differed from those with any level of positivity by a slightly lower incidence of boys, higher mean LDH levels and a 38% (3/8) incidence of relapse, compared to 6.8% (5/74) in all other patients. Histological and immunophenotypic analyses were compatible with a diagnosis of T-LBL. Since we do not know how many of these patients would have been ddPCR negative, this subset needs to be studied in greater detail, once defined by uniform sensitivity MDD quantification.

MRD evaluation in a limited number of patients, predominantly by MCF, confirmed previous reports, with a trend for poor outcome, but failed to detect the majority of relapses and will be difficult to undertake in purely nodal, MDD low/negative patients. Since a disproportionate number of positive results were from the minority of samples assessed by ddPCR, this technique would seem preferable to MFC, as practised here, for MRD assessment. Although it will be possible to perform MRD evaluation in the majority of patients, its value relative to combined oncogenetic/MDD stratification needs should be evaluated within *N/F* defined subgroups, as should its prognostic value compared to imaging evaluation. Whether patients with MDD low/neg. status should be screened for MRD emergence (presuming that tumour immunogenotype/phenotype can be determined from diagnostic tissue) is unclear, but our data illustrate that this is unlikely to be as useful as in T-ALL, somewhat complicating inclusion of T-LBL in MRD-driven T-ALL protocols.

In practical terms, pediatric T-LBLs with *N/F^mut^* respond very well to current standard protocols and MDD status adds little/nothing to their risk assessment. Most would be easily MRD accessible, since 85% (35/41) demonstrated at least 0.01% PB/BM clonal dissemination at diagnosis, but their excellent outcome will make it difficult to demonstrate added value of MRD stratification, and MRD kinetics may differ in T-LBL and T-ALL. Although the small number of relapsing cases precludes definitive assessment, of the 5 *N/F^mut^* patients with MRD evaluation, only two were MRD positive (Table S1).

Amongst *N/F^GL^* patients, MDD < 0.1% status represents a promising means of rapidly identifying patients with a very high relapse risk, as seen in 4/9 patients in the present series (compared to 1/11 patients with MDD > 0.1%). Approximately 80% would be MRD accessible, and the impact of MRD in this small *N/F^GL^*, MDD low/neg. subgroup (9/61, 15%) should be evaluated prospectively, although this will require large patient numbers and universal availability of diagnostic tissue for baseline molecular and immunophenotypic assessment. It will also be important to evaluate the oncogenetic status of these patients, who clearly require early alternative therapy.

## Data Availability

-Central registration of clinical data in Lyon for the French cohort.
-Results and conclusions of the European Intergroup EURO-LB02 trial in children and adolescents with lymphoblastic lymphoma (PMID 28983060)

## Acknowledgements

The authors thank the SFCE and the investigators, the patients and their families from the 24 SFCE-centres involved for providing patient material and data: Anne Lutin, Isabelle Pelier, Nathalie Cheik, Marianna Dapris, Justyna Kanold, Hélène Pacquement, Florent Neumann, Dominique Plantaz, Brigitte Nelken, Charlotte Oudot, Catherine Curtillet, Claudine Schmitt, Marie Laure Couec, Maryline Poiree, Thiery Leblanc, Arnaud Petit, Frederic Millot, Gregory Guimard, Virginie Gandmer, Nimrod Buchbinder, Audrey David.

## This study was supported by

The Necker LBL RELYE (Réseau des lymphomes de l’Enfant) tissue bank is supported by IMAGINE for Margo via the SFCE, the Institut National du Cancer and the CARPEM SIRIC. Research support was provided by the League National de Recherche sur le Cancer (Enfants, Adolescents et Cancer), by Enfants et Santé and by the SFCE. The Trousseau team was supported by the Recherche en Hémopathies Malignes de l’Enfant (RHME).

## Authorship Contributions

ATr, NG and EM conceived the study and oversaw the project; NG, NA, CR, AP, KM, CCC and YB provided study materials or patients; ATr, AP, CA, ATo, LL, VA and EM performed analyses; ATr, LL, VA, NG, CJ, AP and EM collected and assembled data; ATr performed statistical analysis; ATr, AP, CA, ATo, LL, VA and EM interpreted data; ATr, AP, NG and EM wrote the manuscript; all authors approved the manuscript.

## Supplementary data

**Figure S1:**
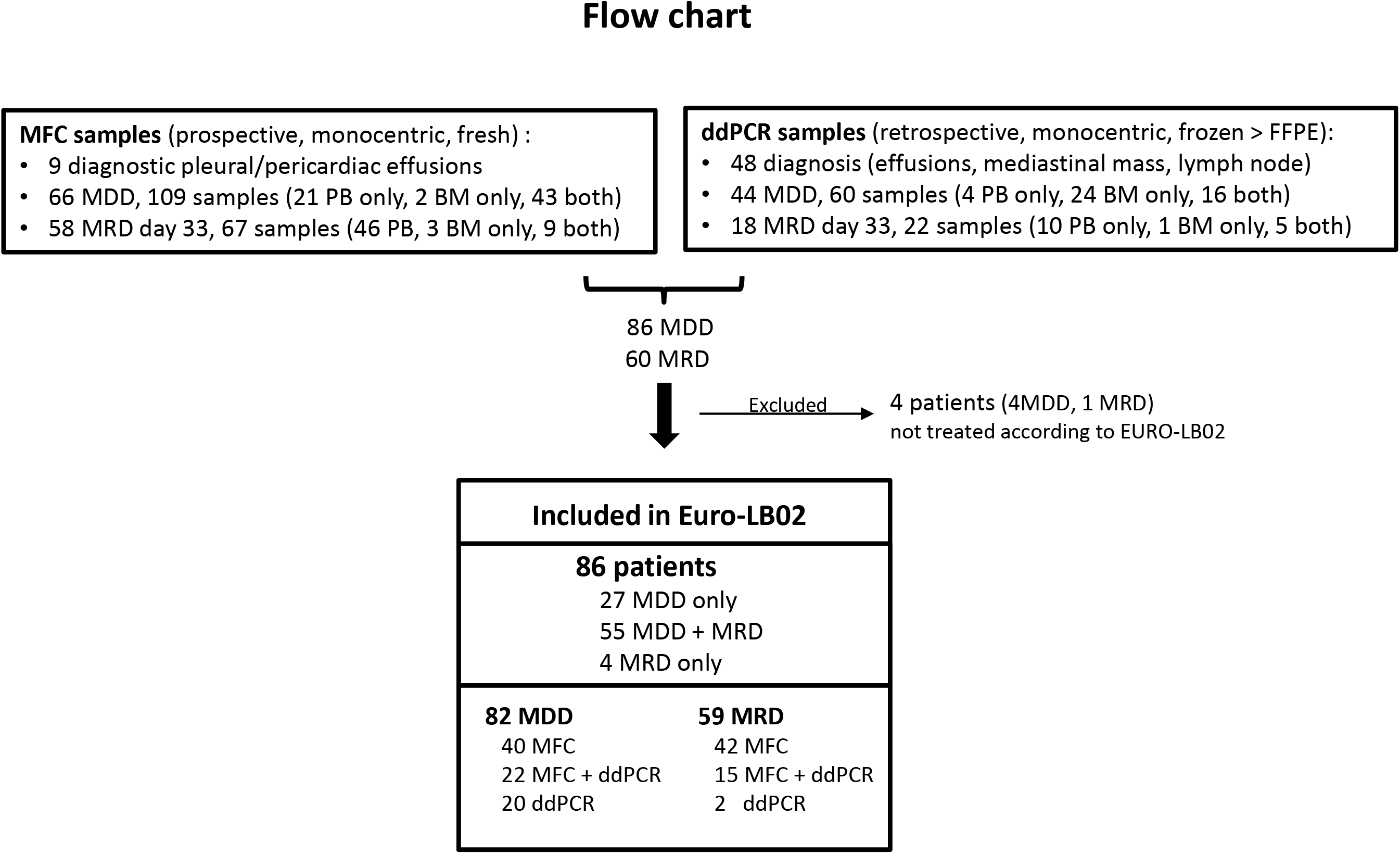
Flow Chart of MDD/MRD evaluation in pediatric T-LBL.

**Figure S2:**
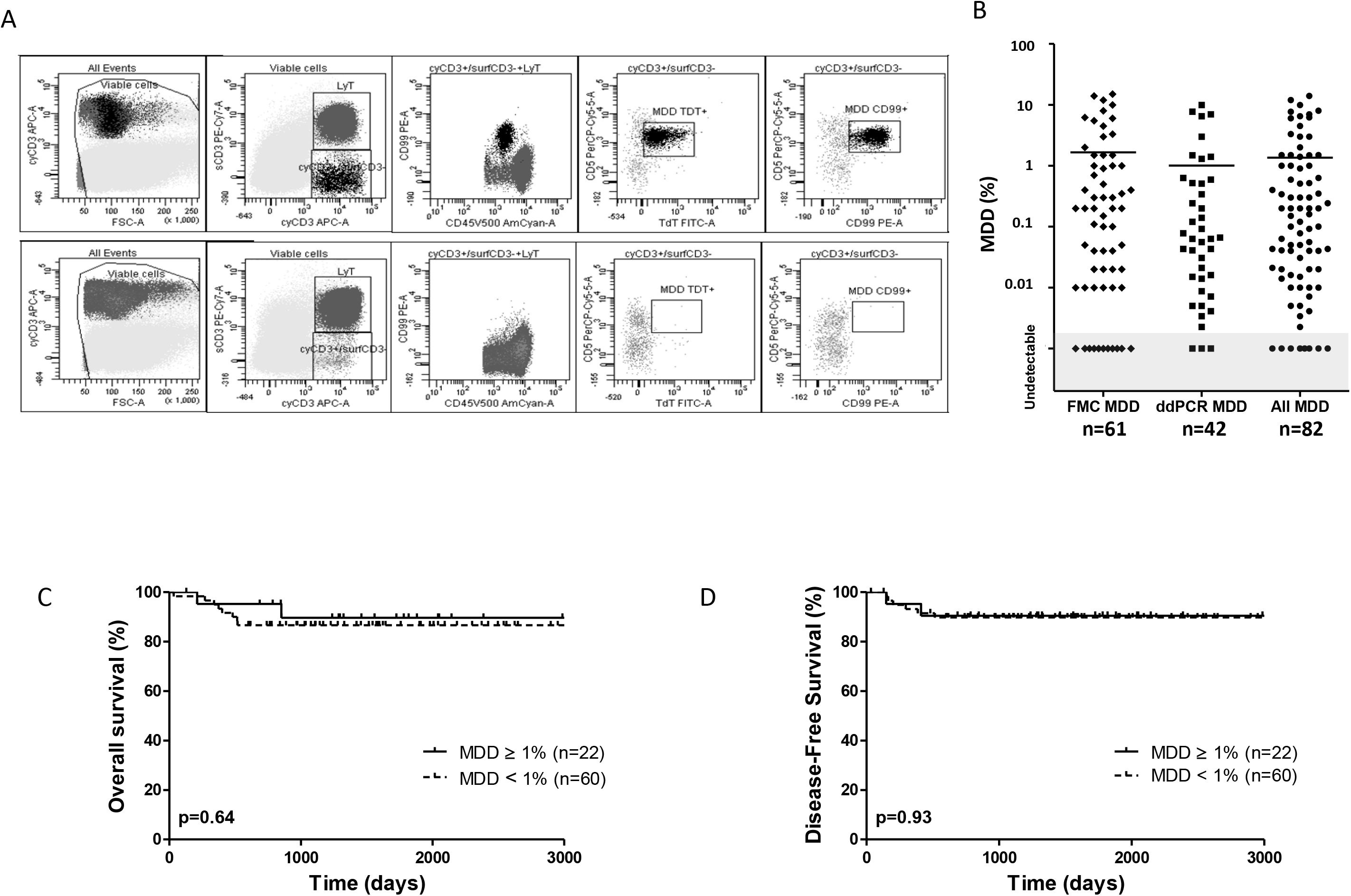
MDD quantification assays. (A) Flow cytometry gating strategy for MDD using FACS Canto II (BD Biosciences) and Diva software. Cells were first gated on viable cells, than CyCD3+/SSC, surf CD3+/SSC, Cy CD3+/surf CD3-. From this last population, blasts were progressively gated by the expression of CD5, TdT and CD99. Back-gated blasts/MDD are stained black. MRD level = events MDD/events of total viable nucleated cells MDD LAIP (CyCD3+/surfCD3-CD99+TdT+). Top: MDD positive at 0.4%, bottom: MDD undetectable. Antibodies used included: TdT (clone HT6, Dako), CD99 (clone TU12, Beckman Coulter, BC), CD2 (clone S5.2, Becton Dickinson, BD), CD5 (clone L17F12, BD), CD7 (M-T701, BD), CD1a (clone BL6, BD), CD4 (clone RPA-T4, BD), CD8 (clone SK1, BD), CD34 (clone 581, BD), CD38 (clone HB7, BD), TCR αβ (clone IP26A, BC), TCRλδ (clone IMMU510, BC), CD56 (clone NCAM16.2, BD), CD45 (clone HI30, Becton Dickinson), surfaceCD3 (clone SK7, BD), cyCD3 (clone UCHT1, BC), CD335/NKP46 (clone 9E2, BD), CD45RA (clone HI100, BD), CD13 (clone L138, BD), CD33 (clone P67.6, BD), CD117 (clone 104D2, BD), CD19 (BD). Cytoplasmic CD3 and nuclear TdT labeling were performed after IntraStain (Dako, Glostrup, Denmark) permeabilisation. (B). Continuum of MDD by MFC, ddPCR or both. The single below quantitative level (BQL) positive result by ddPCR was arbitrarily shown at 0.001% (10^−5^). All MDD refers to samples, with the higher value retained for samples with both ddPCR and MFC analysis. (C) Overall Survival with a MDD 1% threshold. (D) Disease-Free Survival with a MDD 1% threshold.

**Figure S3:**
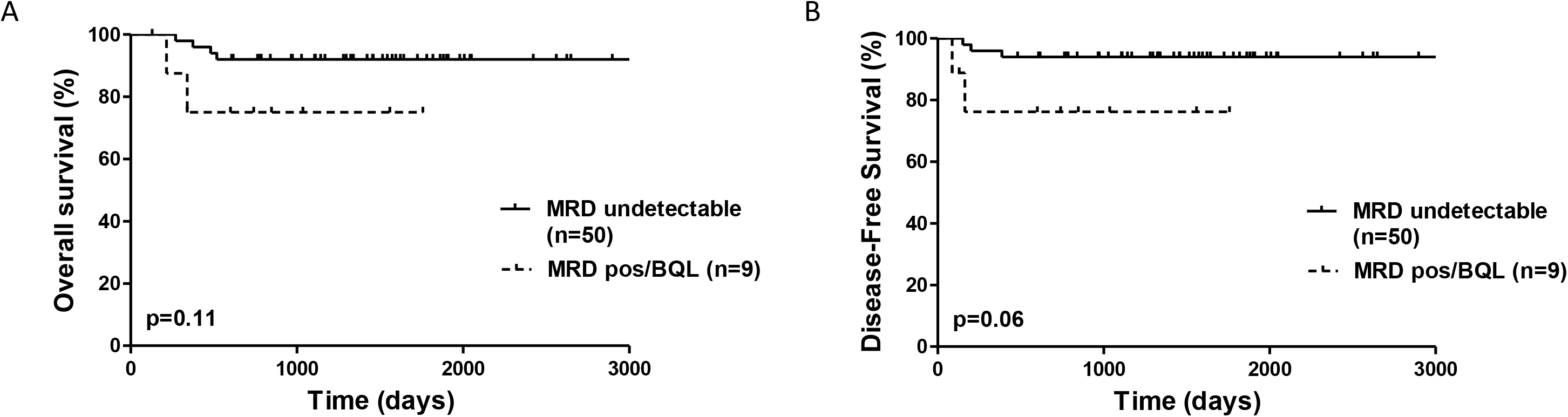
MRD analysis in 59 pediatric T-LBL patients. (A) Overall survival according to MRD analysis in blood or bone marrow at end of induction.(B) Disease free survival according to MRD analysis in blood or bone marrow at end of induction.

**Table S1:** Clinical and biological characteristics of the 9 relapsing and/or those with MDD negativity. ND; Not Done. BQL; Below quantitative level by ddPCR. *Pt. 6239 was not included in the prognostic analyses including N/F status, since the oncogenic status was only performed at relapse.

| Protocol number | Age | Sex | Stage | LDH (xN) | BM | SNC | Relapse | Alive | N/F mutation | MDD value | MDD assay | MRD value | MRD assay |
| --- | --- | --- | --- | --- | --- | --- | --- | --- | --- | --- | --- | --- | --- |
| 6282 | 15.5 | M | IV | 0.9 | Yes | No | Yes | No | Yes | 12% | MFC | ND | ND |
| 6066 | 12.2 | M | II | 1 | No | No | Yes | No | No | 1% | ddPCR | ND | ND |
| 6259 | 12.7 | M | III | 1.4 | No | No | Yes | No | No | 0.02% | MFC + ddPCR | Undetectable | MFC + ddPCR |
| 6031 | 2.9 | M | III | 1 | No | No | Yes | No | Yes | 0.02% | ddPCR | ND | ND |
| 6180 | 16.9 | F | III | 5.9 | No | No | Yes | No | No | 0.003% | MFC + ddPCR | BQL | MFC + ddPCR |
| 6163 | 10.3 | M | III | 1.7 | No | No | Yes | No | No | Undetectable | MFC | ND | ND |
| 6239 | 15.5 | F | III | 2.2 | No | No | Yes | No | ND (No at rel. at 5 m)* | Undetectable | MFC | Undetectable | MFC |
| 6265 | 6.3 | M | III | 15.2 | No | No | Yes | No | No | Undetectable | MFC + ddPCR | Undetectable | MFC |
| 6114 | 16.1 | M | III | 2.9 | No | No | Yes | No | No | ND | ND | 2.00E-04 | MFC |
| 6235 | 5.1 | F | III | 4.1 | No | No | No | Yes | Yes | Undetectable | ddPCR | ND | ND |
| 6284 | 10.3 | F | I | 0.9 | No | No | No | Yes | Yes | Undetectable | MFC + ddPCR | Undetectable | MFC + ddPCR |
| 6184 | 4.3 | M | III | 1.6 | No | No | No | Yes | ND | Undetectable | MFC | Undetectable | MFC |
| 6207 | 15.9 | M | III | 2.4 | No | No | No | Yes | ND | Undetectable | MFC | Undetectable | MFC |
| 6220 | 9.2 | M | III | 1.5 | No | No | No | Yes | ND | Undetectable | MFC | Undetectable | MFC |

